# Analysis of Corticosteroids in Immune Checkpoint Inhibitors (ICE) Induced Myocarditis – A Systematic Review of 352 Screened Articles

**DOI:** 10.1101/2021.11.07.21266031

**Authors:** Mona Sheikh, Saumil Patel, Shavy Nagpal, Zeynep Yulkselen, Samina Zahid, Vivek Jha, Diana F Sánchez-Velazco

**Affiliations:** Division of Clinical and Translational Research, Larkin Community Hospital, South Miami, Florida; University of North Texas Health Science Center, School of Public Health, Fort Worth, Texas; University Of Massachusetts, Massachusetts; Cayetano Heredia Peruvian University, Lima, Perú

**Keywords:** ICI-induced myocarditis, ICI, treatment, diagnosis, corticosteroid

## Abstract

**Introduction:** Immune Checkpoint Inhibitors (ICI) are used as a single agent or as a combination therapies for early or late-stage malignancies. The common malignancies that ICI targets include the following: melanoma, lung cancer, renal cell carcinoma, and hematological malignancies such as Hodgkin’s lymphoma. ICI use is associated with many immune-related adverse events, and ICI-induced myocarditis is one of the rare and most severe AE with a high mortality rate. There are no consensus evidence-based treatment guideline; the expert recommendation is to use high-dose steroids. We aim in this review to assess the effectiveness of steroids in treating ICI-induced myocarditis.

**Methods:** We searched the following database Pubmed, Scopus, Cinhale, and Google Scholar, using the following keywords: ICI-induced myocarditis, treatment, steroid. We included articles in the English language, case reports, case series, and published in the last five years.

**Results:** 352 articles were screened using PRISMA guidelines. After excluding the articles that were duplicate, irrelevant, and did not meet inclusion criteria, 35 articles with a total number of 50 patients were included. All patients treated with ICI either as a single or combination regimen. The onset of symptoms post initiation varied from one day to a year. 46 out of the 50 cases received high doses of Intravenous steroids as a loading dose followed by an oral or intravenous maintenance dose. Out of 50 patients 14 patients (28 %) died but 34 (68 %) patients survived, and 2 (4 %) patients data were not available. The mean age of the patients was 66.31 ± 14.071 (range 23-88 years), 29 were male (58%), 21 were female (42%). Most of the cases were from the USA (42%), followed by Australia (20 %), Japan (14%), Germany, France, and China (4%), Switzerland, Canada, and Spain (2%), and for (6%) cases. A total of 23 patients had cardiovascular comorbidities (46%), which were HTN (14 patients, 60.87%), hyperlipidemia (5 patients, 21.73%), and less than 1 % of patients had myocardial ischemia, congestive heart failure, atrial fibrillation, and peripheral vascular disease. While 26 patients (52%) had normal basal cardiac status.

**Conclusion:** Our results showed that high doses of steroids were effective in controlling cardiac myocyte inflammation and mortality by 28%. Race was not included in the analysis as it was not reported. More in – depth studies are needed to provide a broader representation of steroids in myocarditis.

## Introduction

### Understanding Immune checkpoint inhibitors

Immune Checkpoint inhibitors are monoclonal antibodies (IgG), which are produced by using hybridoma cells. [1] The monoclonal antibody binds to the regulatory receptor on T-cell lymphocyte named programmed cell death receptor 1 (PD-1), programmed cell death ligand 1 (PD-L1), and cytotoxic T-lymphocyte-associated protein 4 (CTLA-4). [2] The normal non-neoplastic cell expresses ligands that bind to these regulatory receptors and prevent them from attacking the normally functioning cell. The tumor cells overexpress PDL-1which helps them to escape destruction by the immune cells. [2]

The monoclonal antibody binds to the regulatory receptors PD-1 (Nivolumab, Pembrolizumab, Cemiplimab), PD-L1 (Avelumab, Durvalumab, Atezolizumab), and CTLA-4 (Ipilimumab), the binding enables the immune system to attack and kill the cancer cells. [3] ICIs were reported to induce many immune-related adverse events (irAE) including, myositis, hepatitis, myasthenia gravis, and myocarditis. [4] Treatment with ICIs does not only affect the tumor cells but also the non-tumor cells as well. ICI use was found to cause an influx of lymphocytes and macrophages into the myocardium, which ultimately causes myocarditis. [5]

Based on the ICI mechanism of action, there is a theory that the myocardium expresses an antigen that is similar to the tumor cell antigen; therefore, it becomes a T-Lymphocyte target, and lymphocytes infiltrate the myocardium. [4] ICI-induced myocarditis is a rare irAE that presents with non-specific symptoms and is associated with a high mortality rate. [6] The clinical presentation of myocarditis varies widely, ranging from chest pain, fever, dyspnea, increased heart rate, and general body aches. [7, 8] Diagnosis confirmed with endomyocardial biopsy. [8] However, since this is an invasive test, initial investigations include cardiac labs such as Creatine kinase (CK), Troponin I (TnI), and Troponin T (TnT), echocardiogram, and cardiac MRI. [7,9] Myocarditis predominantly affects males as compared to females with individuals above the age of 40 years. [8]

There are no randomized controlled trials conducted to have evidence-based treatment guidelines for ICI-induced myocarditis treatment. The guidelines recommend discontinuation of the ICI and giving high-dose steroids, which are expert experience and agreements. We are aiming in this systematic review to evaluate the effectiveness of steroids and obtain a valid conclusion.

## Methods

### Search Strategy

This systematic review was conducted according to the Preferred Reporting Items for Systematic Reviews and Meta-Analyses (PRISMA) guidelines. An extensive literature search was performed in PubMed, Scopus, and CINAHL for case reports and case series. The following terms were used in MeSH and free-text searches: myocarditis, cancer, corticosteroid, steroid, immune checkpoint inhibitor, immune checkpoint blockade, checkpoint inhibitor, immune-related adverse events, immunotherapy, adverse effect, immunosuppressive, glucocorticoid, cyclosporine, mycophenolate mofetil, azathioprine, prednisone, methylprednisolone, infliximab, anti-thymocyte globulin, drug-induced myocarditis. The “related articles” function of PubMed was used for broadening the search. We attempted to identify additional studies by searching the reference lists of selected articles.

#### Inclusion Criteria

We included all case reports and case series that met the following criteria:

⍰ Studies focused on myocarditis induced by Immune checkpoint inhibitors and received steroid or non-steroid as a treatment.
⍰ Myocarditis is diagnosed by biopsy or CMRI (cardiac magnetic resonance imaging) or autopsy l1 Time interval between years 2016-2020
⍰ Articles published in English in a peer-reviewed journal
⍰ Participants are at least 18 years old or above.

#### Exclusion Criteria

We excluded articles with the following criteria:

⍰ patients aged < 18 years
⍰ patients did not receive myocarditis treatment or the treatment not reported
⍰ myocarditis is not induced by immune checkpoint inhibitors
⍰ the diagnosis is not confirmed by biopsy or cardiac MRI
⍰ non-English articles.

### Study Selection

All case reports and case series studies that evaluated the efficacy of steroid or non-steroid treatment in ICI-induced myocarditis were downloaded into Covidence from search engines. During the first step of this literature search, title and abstract screening were done by two independent reviewers using Covidence. A third independent reviewer resolved any conflict that arose. After completing the title and abstract screening, the full-text of eligible studies was screened in detail by two independent reviewers according to the inclusion and exclusion criteria set before. Any conflict was resolved by a third independent reviewer. Eventually, studies were selected for data extraction. Two independent reviewers performed the screening of each study and any type of discrepancies were resolved through discussions until they reached a consensus. [Figure 1]

**Figure 1:**
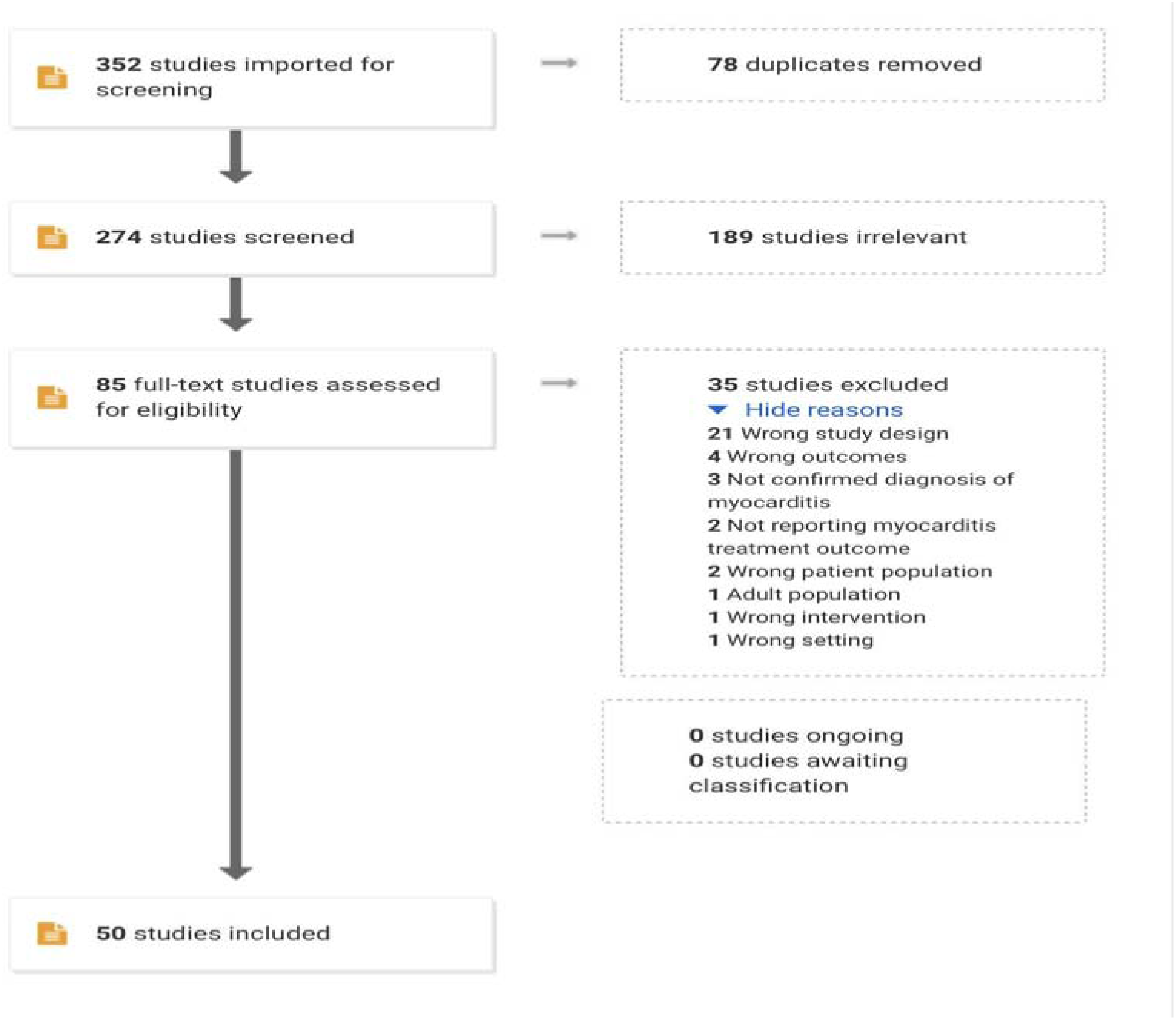
Prisma chart for study acquisition.

### Data Extraction

For all included studies, six review authors independently extracted data and summarized characteristics in tables. Authors mainly extracted these characteristics: author name, publication year, patient age and gender, type of cancer, stage of cancer, comorbidity, symptoms, ECG, echocardiogram, lab values, CMR, biopsy, type of myocarditis treatment (initial treatment, secondary treatment), the dose of steroid or other drugs, the outcome of initial and secondary treatment, prognosis, mortality, autopsy.

### Data Analysis

We used IBM SPSS and Microsoft Excel software to run data analysis. Continuous variables are presented as mean ± SD, and categorical variables are presented as absolute numbers and percentages.

### Assessment of risk of bias

JBI critical appraisal checklists for case series were applied for quality and risk of bias assessment. There were eight questions in the checklist against which the quality of the study was evaluated. There were three possible answers to the questions, either yes, no, or unclear. Based on that, the overall appraisal of the studies was determined. [Table 1]

**Table1:** Bias assessment.

|  | First Author | Q1 | Q2 | Q3 | Q4 | Q5 | Q6 | Q7 | Q8 | Overall appraisal |
| --- | --- | --- | --- | --- | --- | --- | --- | --- | --- | --- |
| 1 | Saibil 2019 | yes | yes | yes | yes | yes | yes | unclear | yes | Include |
| 2 | Monge 2018 | yes | Yes | Yes | Yes | Yes | Yes | No | Yes | Include |
| 3 | Guo 2020 | Yes | Yes | Yes | Yes | Yes | Yes | unclear | Yes | Include |
| 4 | Padegimas 2019 | Yes | Yes | Yes | Yes | Yes | Yes | No | Yes | Include |
| 5 | Salem 2019 | No | Yes | Yes | Yes | Yes | Yes | No | Yes | Include |
| 6 | Liu 2020 | Yes | Yes | Yes | Yes | Yes | Yes | unclear | yes | Include |
| 7 | Jain 2018 | Yes | No | Yes | Yes | Yes | Yes | unclear | yes | Include |
| 8 | Semper 2016 | Yes | Yes | Yes | Yes | Yes | Yes | unclear | Yes | Include |
| 9 | **Tadokoro 2016** | Yes | Yes | Yes | Yes | Yes | Yes | *unclear* | Yes | Include |
| 10 | **Ganatra 2018** | Yes | Yes | Yes | Yes | Yes | Yes | No | Yes | Include |
| 11 | **Frigeri 2018** | Yes | Yes | Yes | Yes | Yes | Yes | *unclear* | Yes | Include |
| 12 | **Heinzerling 2016** | Yes | Yes | Yes | Yes | Yes | Yes | *unclear* | Yes | Include |
| 13 | **Gallegos 2019** | Yes | Yes | Yes | Yes | Yes | No | No | Yes | Include |
| 14 | **Johnson 2016** | Yes | Yes | Yes | Yes | Yes | Yes | No | Yes | Include |
| 15 | **Matsui 2020** | Yes | Yes | Yes | Yes | Yes | Yes | No | Yes | Include |
| 16 | **Edahiro 2020** | Yes | Yes | Yes | Yes | Yes | Yes | No | Yes | Include |
| 17 | **Imai 2019** | Yes | Yes | Yes | Yes | Yes | Yes |  | Yes | Include\ |
| 18 | **Norwood 2017** | Yes | Yes | Yes | Yes | Yes | Yes | No | Yes | Include |
| 19 | **Leaver 2020** | Yes | Yes | Yes | Yes | Yes | Yes | No | Yes | Include |
| 20 | **LoongTan 2020** | Yes | Yes | Yes | Yes | Yes | Yes | No | Yes | Include |
| 21 | **Matsuo 2019** | Yes | Yes | Yes | Yes | Yes | Yes | No | Yes | Include |
| 22 | **Chang 2018** | Yes | Yes | Yes | Yes | Yes | Yes | No | Yes | Include |
| 23 | **Martin Huertas 2019** | Yes | No | Yes | Yes | Yes | Yes | No | Yes | Include |
| 24 | **Tay 2017** | Yes | No | Yes | Yes | Yes | Yes | No | Yes | Include |
| 25 | **Lee 2020** | Yes | Yes | Yes | Yes | Yes | Yes | No | Yes | Include |
| 26 | **Hardy 2020** | Yes | Yes | Yes | Yes | Yes | Yes | No | Yes | Include |
| 27 | **Tan 2020** | Yes | Yes | Yes | Yes | Yes | Yes | No | Yes | Include |
| 28 | **Mahmood 2018** | Yes | Yes | Yes | Yes | Yes | Yes | No | Yes | Include |
| 29 | **Lie 2020** | Yes | Yes | Yes | Yes | Yes | Yes | No | Yes | Include |
| 30 | **Mehta 2016** | Yes | Yes | Yes | Yes | Yes | Yes | No | Yes | Include |
| 31 | **Ansari-Gilani 2020** | Yes | Yes | Yes | Yes | Yes | Yes | No | Yes | Include |
| 32 | **Fukasawa** | Yes | Yes | Yes | Yes | Yes | Yes | Unclear | Yes | Include |
| 33 | **Yamaguchi** | Yes | Yes | Yes | Yes | Yes | Yes | Unclear | Yes | Include |
| 34 | **Tajmir-Riahi** | Yes | Yes | Yes | Yes | Yes | Yes | Unclear | Yes | Include |
| 35 | **Arangalage** | Yes | Yes | Yes | Yes | Yes | Yes | No | Yes | Include |

*Bias assessment answer: Yes/No/Unclear/ Not Applicable*
*Overall appraisal: included/excluded/seek further information*
Q1: Were the patient's demographic characteristics clearly described?
Q2: Was the patient's history clearly described and presented as a timeline?
Q3: Was the current clinical condition of the patient on presentation clearly described?
Q4: Were diagnostic tests or assessment methods and the results clearly described?
Q5: Was the intervention(s) or treatment procedure(s) clearly described?
Q6: Was the post-intervention clinical condition clearly described?
Q7: Were adverse events (harms) or unanticipated events identified and described?

## Results

We identified 352 articles through our search from PubMed, Scopus, and CINAHL. From the 35 articles we selected, a total of 50 patients met our inclusion criteria and were analyzed. Of 50 patients, 26 had malignant melanoma (53%), 8 had carcinoma of the lung (16 %), 4 had renal cell carcinoma (8%), 1 had bladder cell carcinoma (2%), other carcinomas account for less than 2% in remaining individuals. [Table 3] Regarding the cancer stage, 35 patients (70%) were in Stage IV cancer, 7 (14%) were in stage III and remained in the lower stage or undetermined.

42 out of 50 patients (84 %) were symptomatic, major presenting cardiovascular symptoms were dyspnea (22 patients, 41.50%), chest pain (9 patients, 18 %), and chest tightness and edema (4 patients, 8 %). On the other hand, 8 out of 50 patients (16 %) were asymptomatic.

All 50 patients who received immune check inhibitors have developed myocarditis. [Table 4] Among them, 32 patients (64 %) received nivolumab, 19 patients (38 %) received Ipilimumab, 2 patients (4 %) received vemurafenib and Atezolizumab, the patients received Pembrolizumab, Temozolomide, durvalumab, tremelimumab as a combination of about drugs. 17 out of the 50 reviewed cases were on a combination regimen of either Nivolumab and Ipilimumab (15 cases, 30%), durvalumab, and tremelimumab (1 case, 2%), and Nivolumab and Pembrolizumab (1 case, 2%).[Table 4]

Out of 50, 47 patients had been diagnosed with either cardiac MRI or cardiac biopsy, and the remaining 3 patients were diagnosed with autopsy reports. The time of the onset of symptoms after initiation of immune check inhibitors ranges from 1st day after initiation of the treatment to 1 year (from 1st dose to the 13th dose). Those who were on combination ICI had earlier symptoms compared to patients on one ICI, 10 out of 17 patients who were on combination regimen presented with their symptoms after the first dose. [13, 18, 18, 24, 27, 25, 36, 37]

Out of 50 patients, 23 patients (46%) had elevated CK, 13 (26 %) patients had elevated CK-MB, 37 (74 %) patients had increased troponin I/T, 13 (26 %) patients had increased BNP level, 7 (14 %) patients had increased other test including AST, ALT, ANA titer, CRP, LDH, interleukin, myoglobin levels. The mean of ejection fraction was 39.56 % ± 17.72 % (13%-73%).

46 out of the 50 cases received high doses of Intravenous steroids as a loading dose followed by an oral or intravenous maintenance dose. Out of 50 patients 14 patients (28 %) died but 34 (68 %) patients survived, and 2 (4 %) patients’ data was not available. In terms of steroid effectiveness, 47.16% of patients improved while 52.83% did not improve completely and needed adding additional regimen, the added treatment infliximab, IgG, mycophenolate, Tacrolimus, plasma exchange, and plasmapheresis.

The causes of death in the 14th mortality were as following myocardial infarction (MI) [4, 18, 23, 33], multiple organ failure (MOF) [18, 36], complete heart block (CHB) [19, 36], ventricular tachycardia (VT) [18, 4], congestive heart failure (CHF) [19, 30], PE/bacteremia [31], hypotension/diaphragmatic paralysis [37], and progressive lung cancer [11]. The follow-up time ranged from 3 days to 26 months. 3 out of the 4 MI deaths occurred within 7 days of hospital admission, VT also within the first couple days of admission, lastly, the 2 deaths of CHB were on the third and 17th days.

## Discussion

Immune checkpoint inhibitors are a cutting-edge, promising therapy for many malignancies. The FDA approved several ICI in the last decade; initial approval for Ipilimumab, nivolumab, Pembrolizumab, Cemiplimab, Avelumab, Durvalumab, and Atezolizumab was on 28 March 2011, 22 December 2014, 4 September 2014, 28 September 2018, 18 November 2015, 17 February 2016, and 18 May 2016, respectively. [2] They were approved as a single agent or as a combination regimen for advanced or early stages of the following tumor melanoma, renal cell carcinoma (RCC), squamous non-small-cell lung cancer (NSCLC), urothelial cancer such bladder urothelial cancer, colorectal cancer, hepatocellular carcinoma (HCC), head and neck squamous cell carcinoma (HNSCC), cutaneous squamous cell carcinoma (CSCC), Hodgkin’s lymphoma, Merkel Cell carcinoma (MCC), cervical and breast cancer. [2] [Table 2]

**Table 2:**
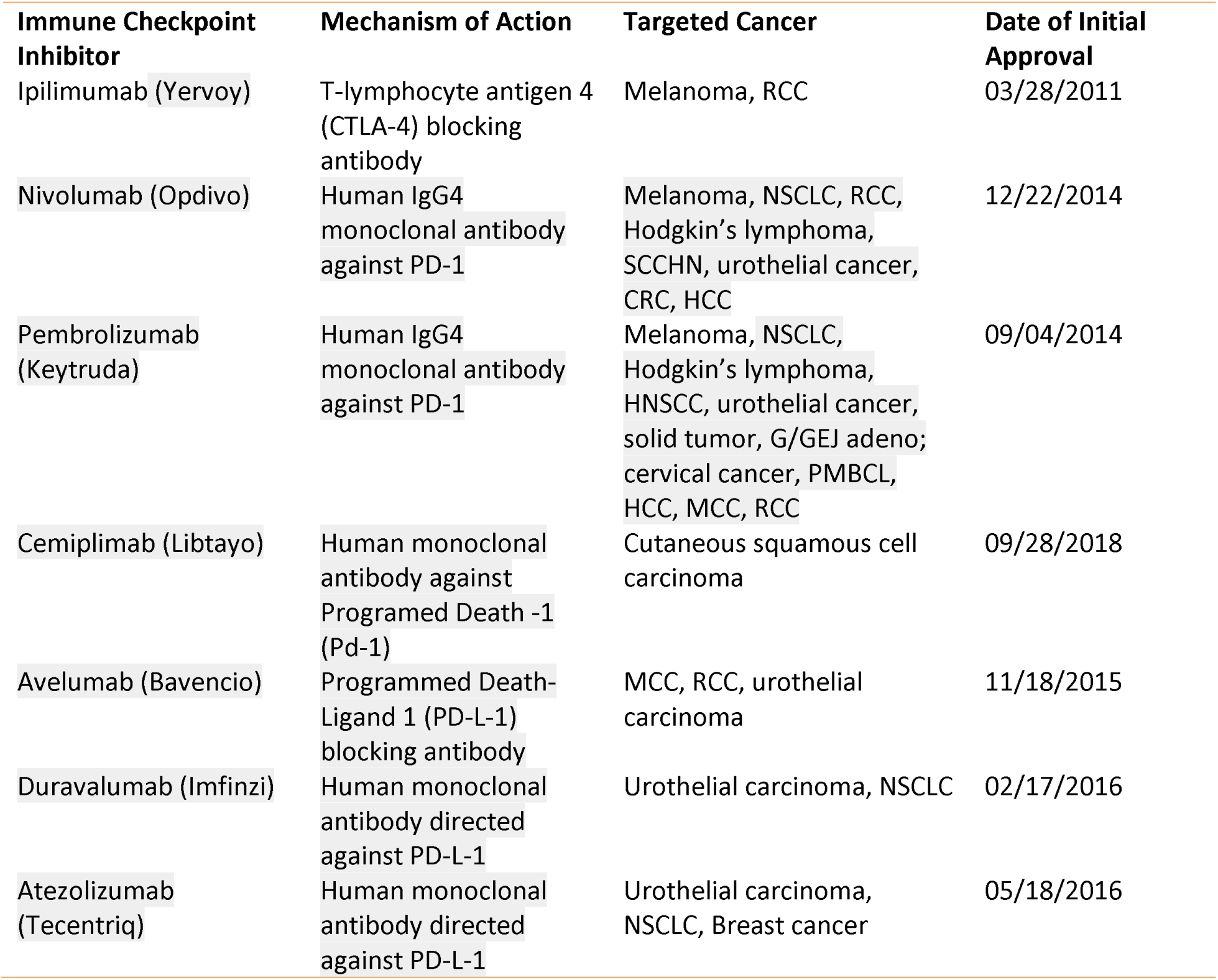
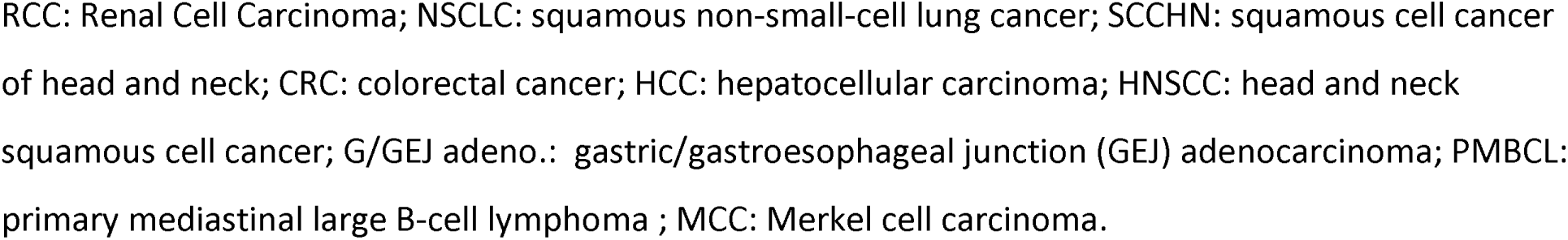
ICI mechanism of action, targeted cancer and date of approval.

| Immune Checkpoint Inhibitor | Mechanism of Action | Targeted Cancer | Date of Initial Approval |
| --- | --- | --- | --- |
| Ipilimumab (Yervoy) | T-lymphocyte antigen 4 (CTLA-4) blocking antibody | Melanoma, RCC | 03/28/2011 |
| Nivolumab (Opdivo) | Human IgG4 monoclonal antibody against PD-1 | Melanoma, NSCLC, RCC, Hodgkin's lymphoma, SCCHN, urothelial cancer, CRC, HCC | 12/22/2014 |
| Pembrolizumab (Keytruda) | Human IgG4 monoclonal antibody against PD-1 | Melanoma, NSCLC, Hodgkin's lymphoma, HNSCC, urothelial cancer, solid tumor, G/GEJ adeno; cervical cancer, PMBCL, HCC, MCC, RCC | 09/04/2014 |
| Cemiplimab (Libtayo) | Human monoclonal antibody against Programmed Death -1 (Pd-1) | Cutaneous squamous cell carcinoma | 09/28/2018 |
| Avelumab (Bavencio) | Programmed Death-Ligand 1 (PD-L-1) blocking antibody | MCC, RCC, urothelial carcinoma | 11/18/2015 |
| Duravalumab (Imfinzi) | Human monoclonal antibody directed against PD-L-1 | Urothelial carcinoma, NSCLC | 02/17/2016 |
| Atezolizumab (Tecentriq) | Human monoclonal antibody directed against PD-L-1 | Urothelial carcinoma, NSCLC, Breast cancer | 05/18/2016 |

RCC: Renal Cell Carcinoma; NSCLC: squamous non-small-cell lung cancer; SCCHN: squamous cell cancer of head and neck; CRC: colorectal cancer; HCC: hepatocellular carcinoma; HNSCC: head and neck squamous cell cancer; G/GEJ adeno.: gastric/gastroesophageal junction (GEJ) adenocarcinoma; PMBCL: primary mediastinal large B-cell lymphoma ; MCC: Merkel cell carcinoma.

**Table 3:**
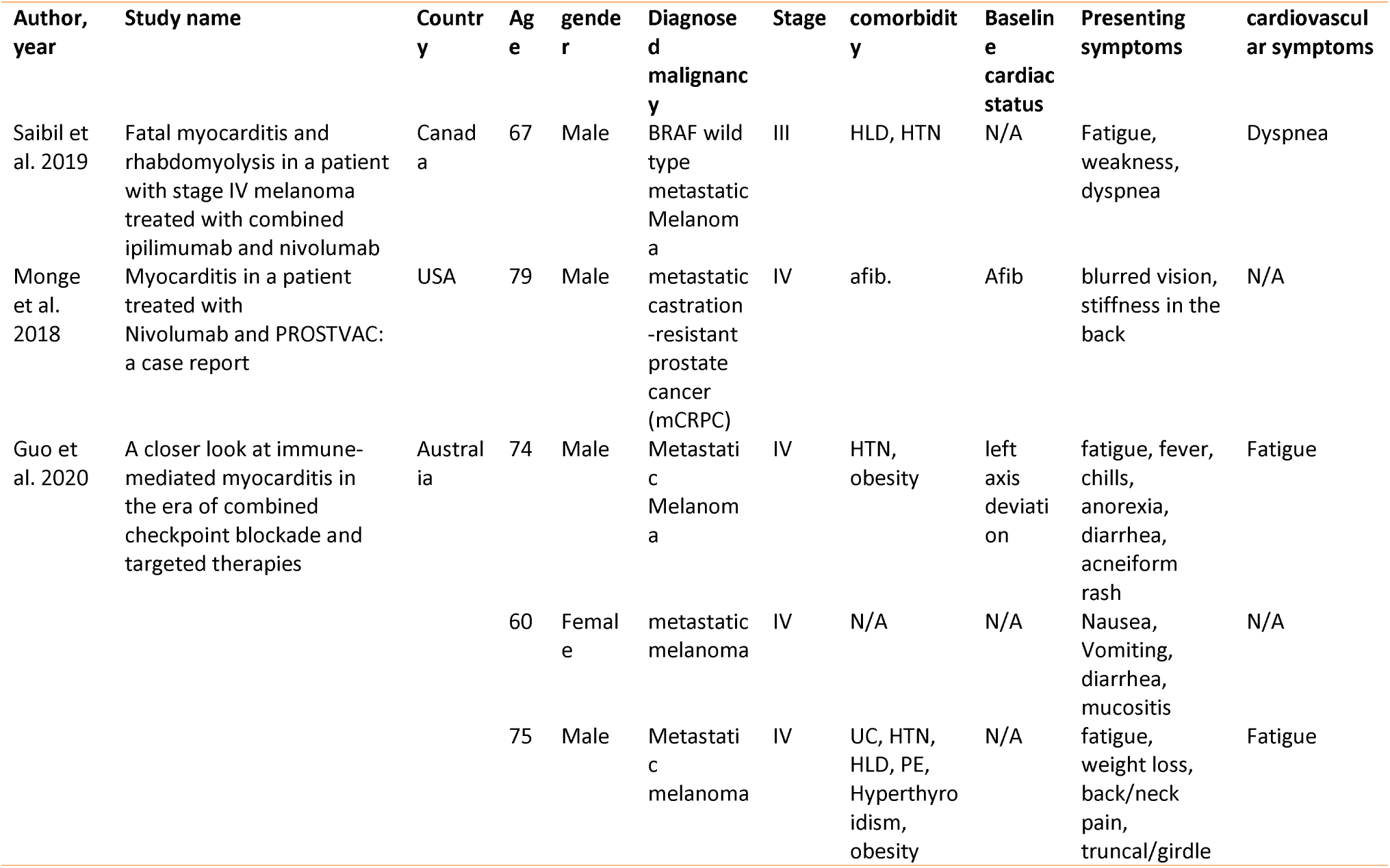
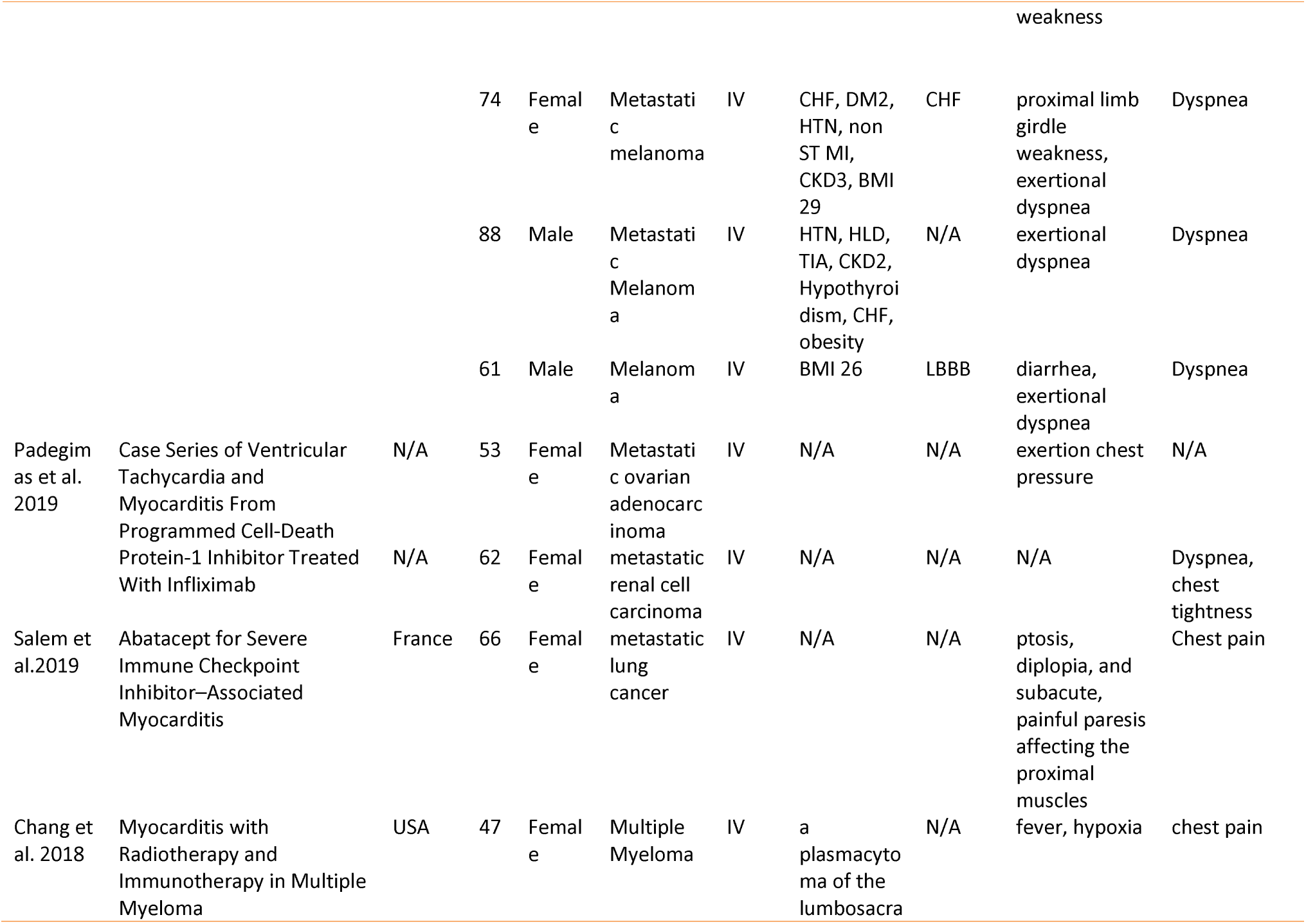
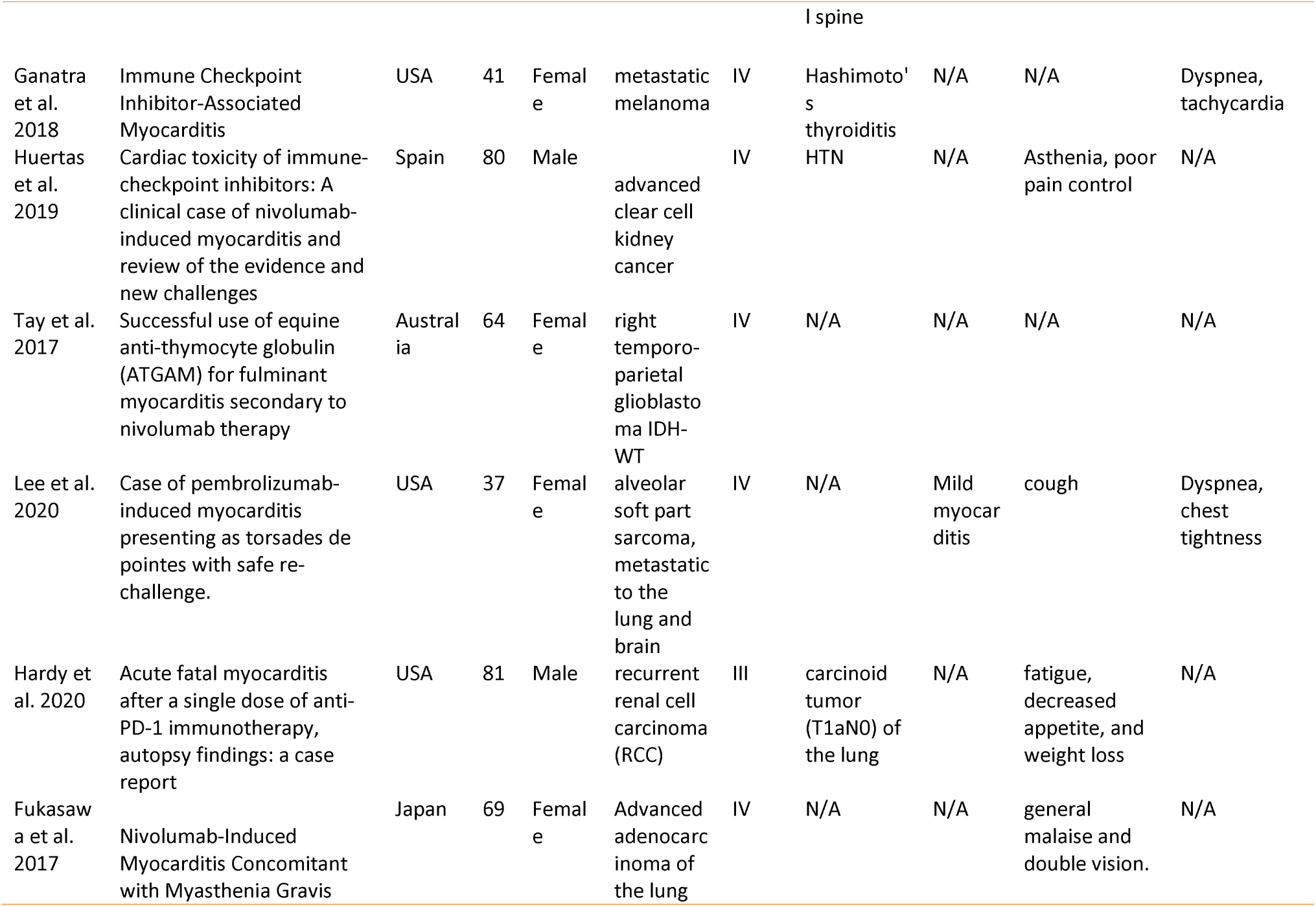
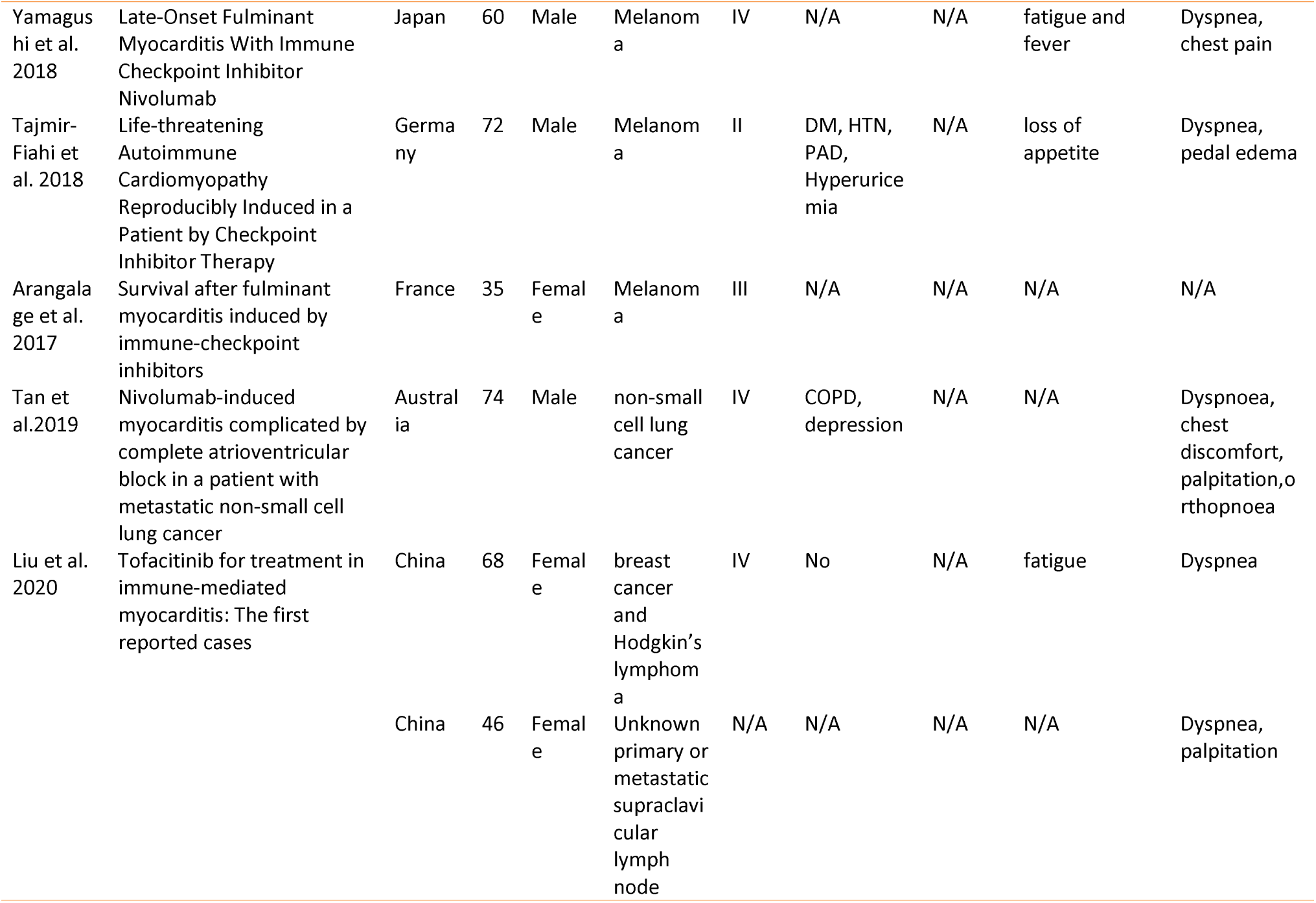
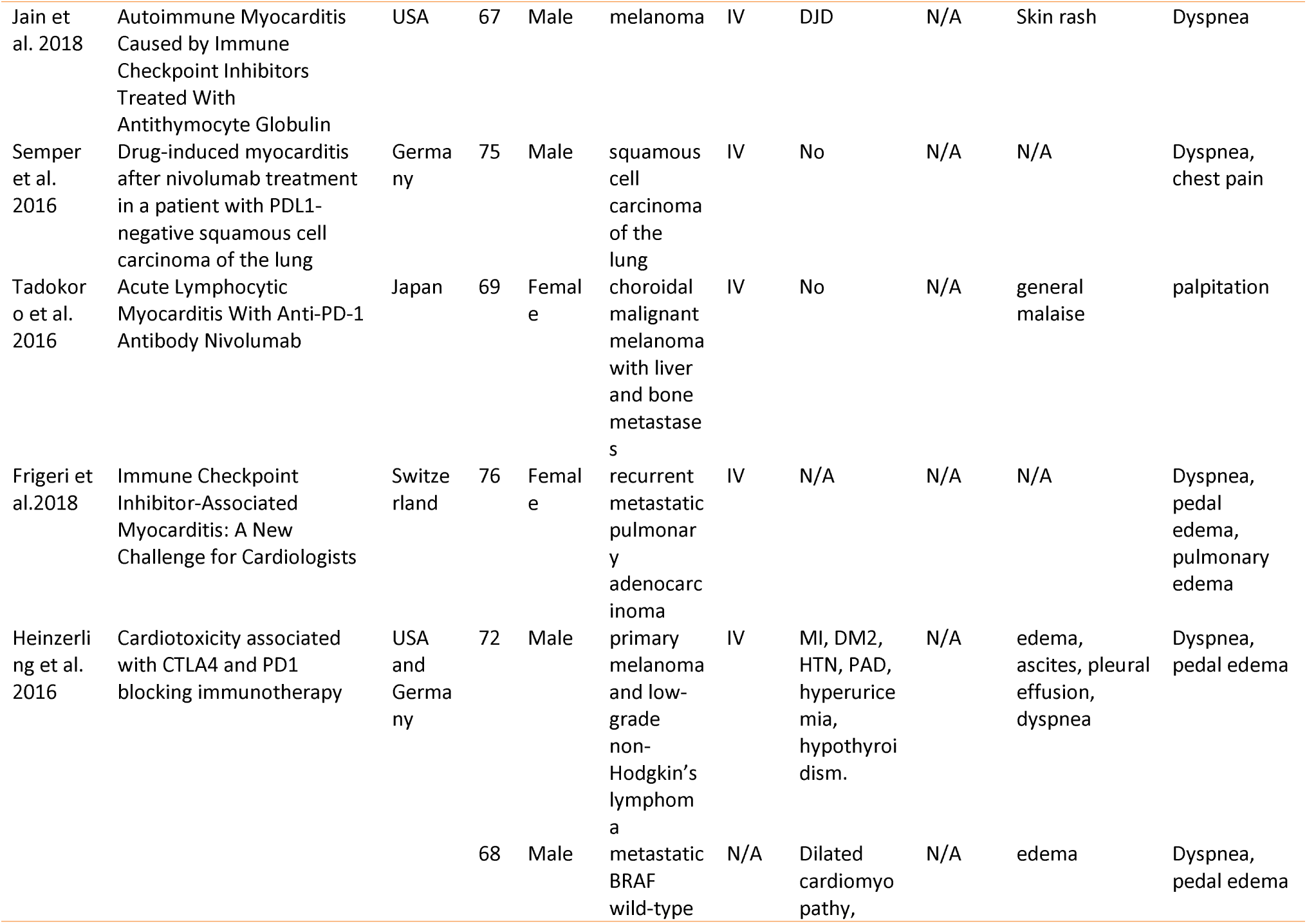
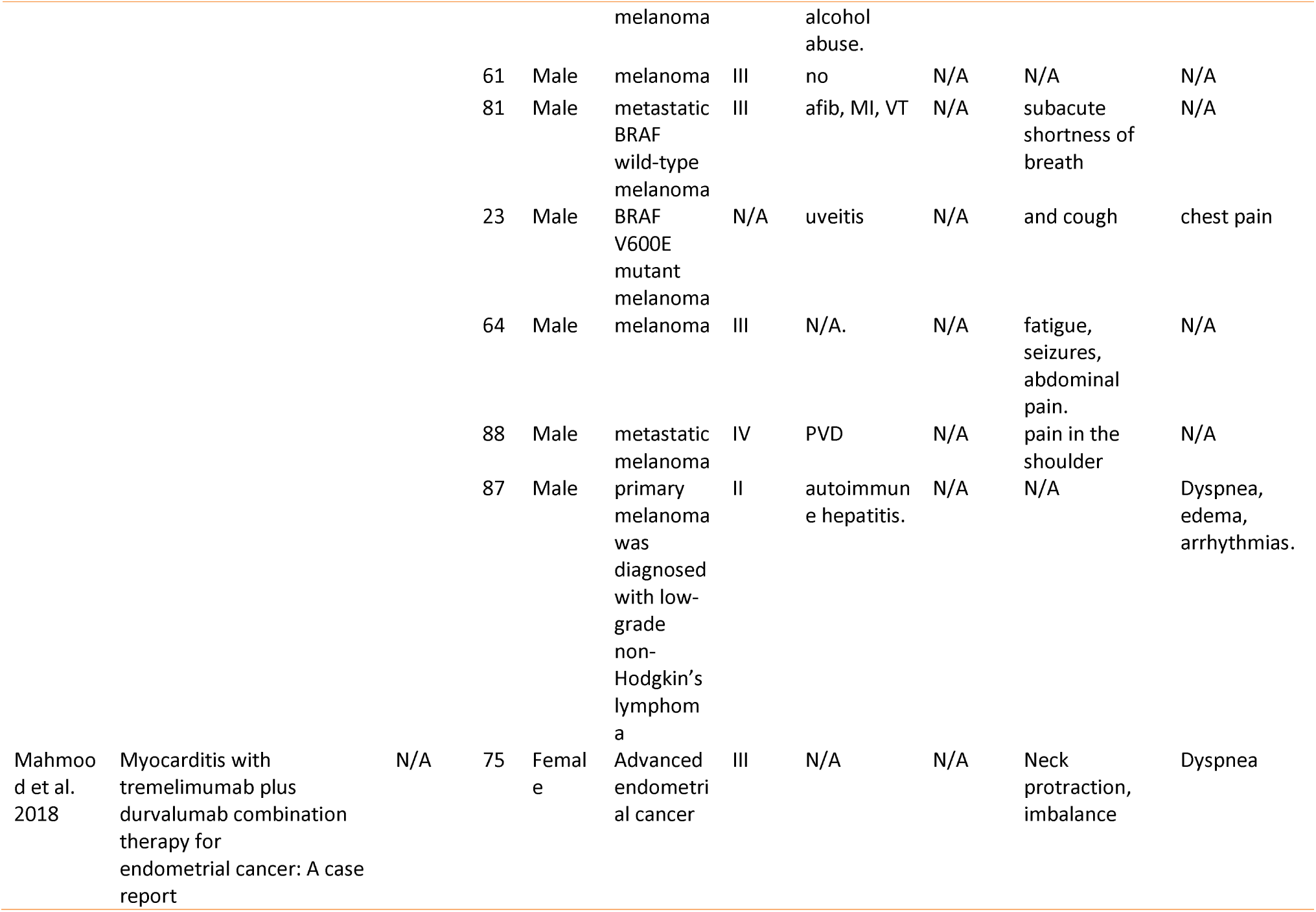
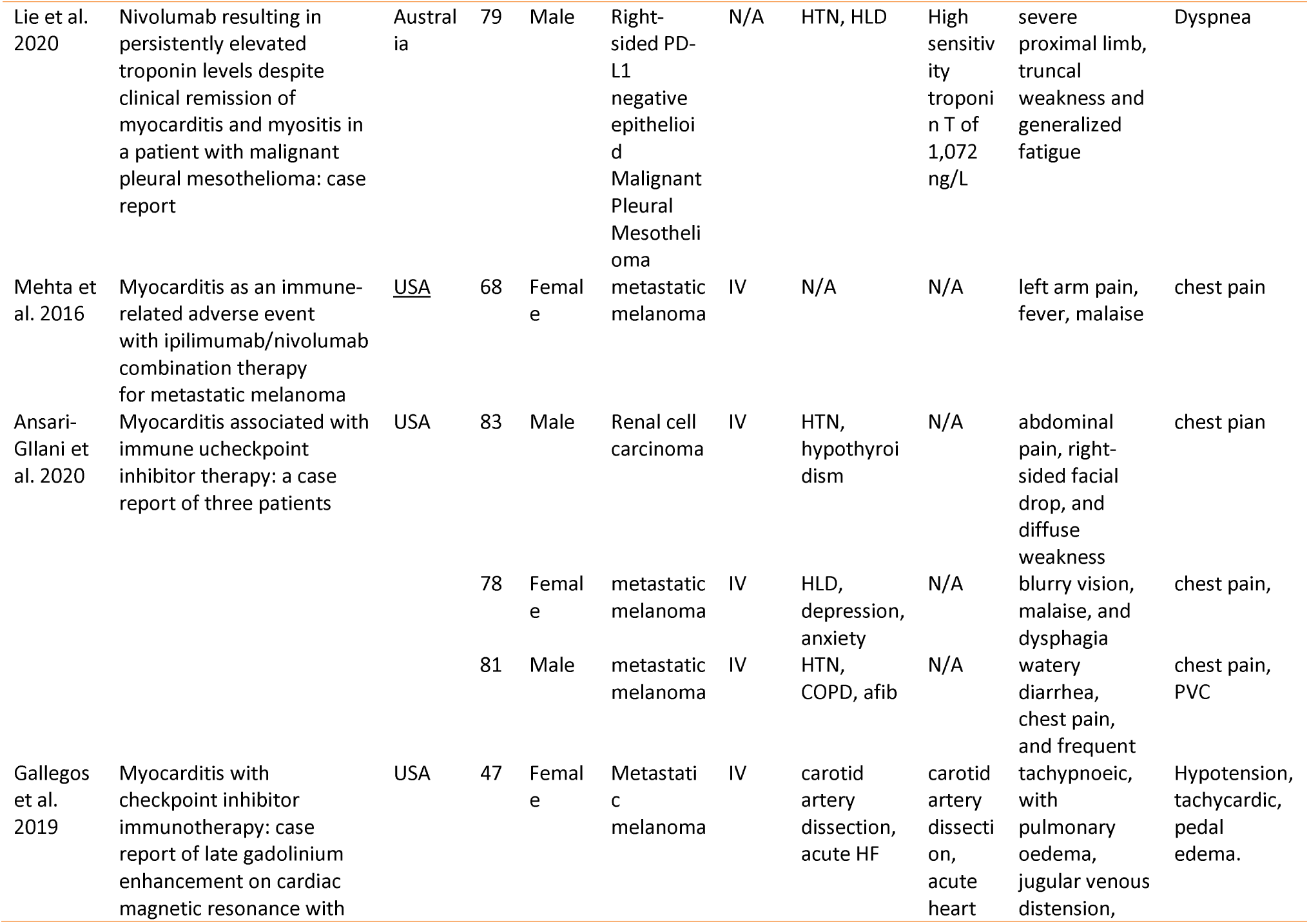
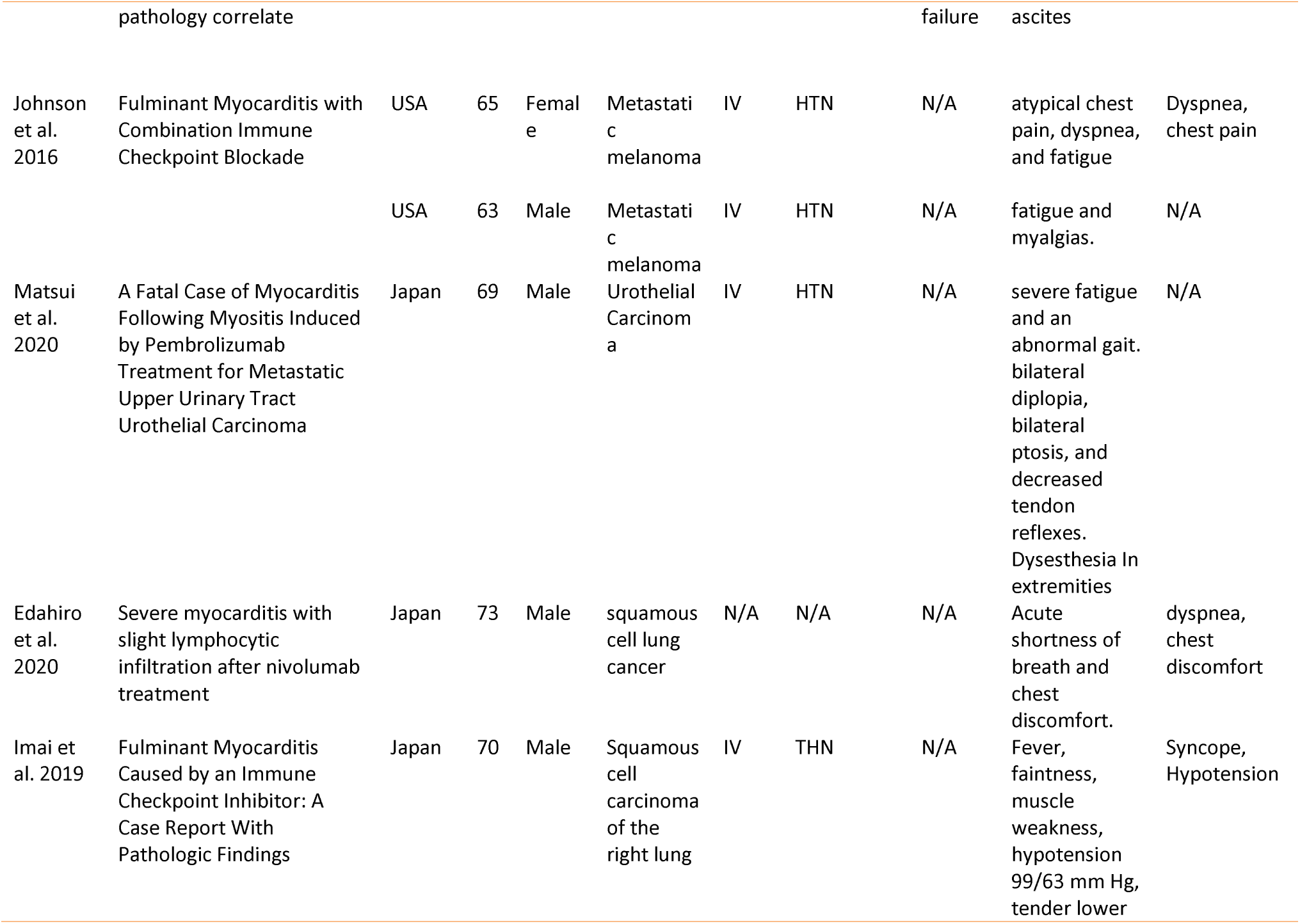
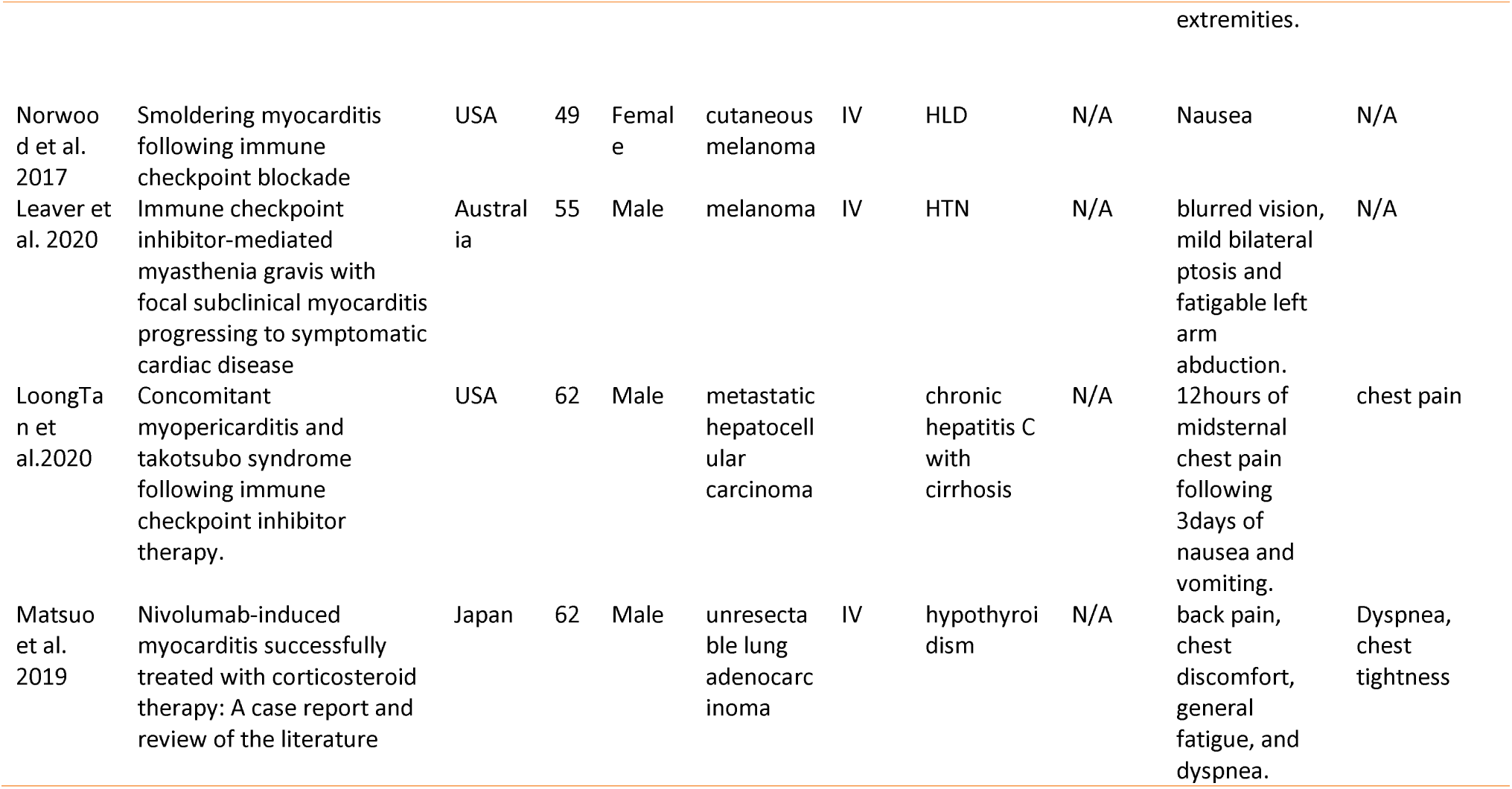
Study details and demographic.

**Table 4.**
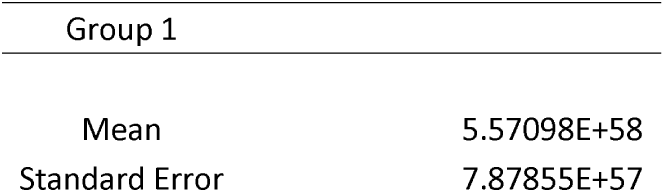
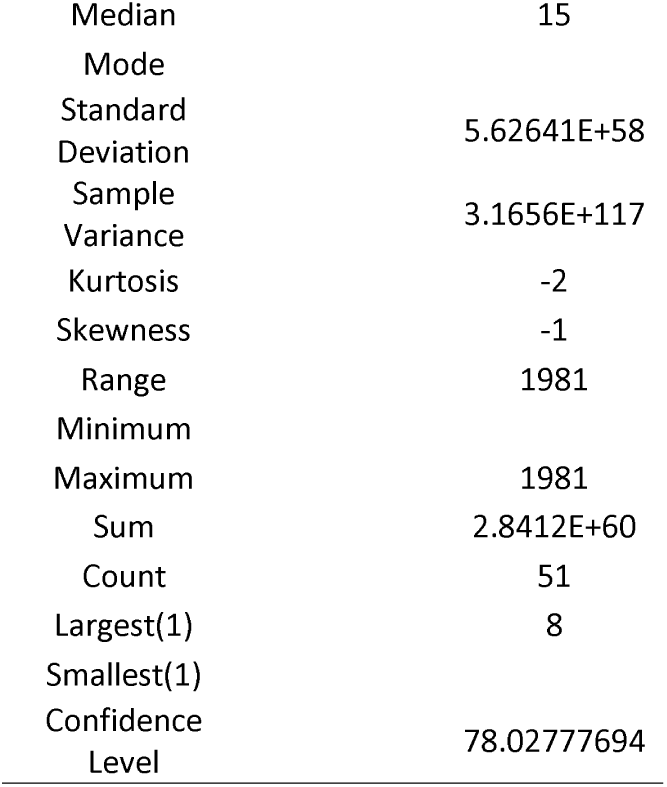
Statistical analysis.

ICI-induced myocarditis is one of the rare and severe forms of irAE of ICI with a high mortality rate. The myocarditis was underestimated during the ICI clinical trials due to the low incidence and vague clinical presentation. For example, in the Nivolumab clinical trial, the most common adverse events were fatigue (33%), nausea (14%), and pruritus (14%); and no cardiac adverse event or death was reported due to the drug toxicity. [4]

Our result showed 81.13% of the patients with ICI-induced myocarditis had cardiovascular symptoms, which were dyspnea (41.50%), chest pain (16.98%), and chest tightness and edema (7.54%). Also, the common general presenting symptoms were fatigue, myalgia, general discomfort, or nausea, which are similar to what is reported in other studies.[7, 8] In addition to the non-specificity of clinical presentation which poses a challenge in the diagnosis, the median reporting time varies which adds another hurdle to diagnose a case of ICI-induced myocarditis. Our results showed the median time onset of symptoms after initiation of the therapy was 79.50 days, 16 days reported by Matznel et al, and 6 months reported in Dolldile et al cohort retrospective study and a year in Gallegos et al. [10, 17, 44] Hence, clinicians should have a high index of suspicion when managing patients on an ICI treatment regimen even if the treatment was initiated over 6 months or a year ago and should be aware of the ICI cardiac adverse events and the need for prolonged cardiac monitoring.

The diagnosis of ICI Induced-myocarditis is predictive of abnormal values in cardiac enzymes such as troponin, ECG, and echocardiogram. The confirmatory tests are cardiac MRI (CMRI) and endomyocardial biopsy. [42, 43] Endomyocardial biopsy is the gold standard test to diagnose ICI-induced myocarditis, however, it is an invasive procedure with high false negative probability due to patchy distribution of the inflammatory lesions and probability of missing to sample the inflamed region of the heart. The microscopic findings are commonly T-cell lymphocyte predominant infiltration and fibrosis which are similar to graft rejection. [18] Cardiac MRI, a noninvasive, detects the macroscopic changes in the heart with late gadolinium enhancement that reflects the edema in the myocyte secondary to inflammation. Cardiac MRI also is used to monitor the progression of the disease and response to therapy. [42] ECG abnormalities vary and are non-specific, including tachycardia, bradyarrhythmia, heart block, atrial and ventricular tachyarrhythmia. We found complete heart block and ventricular tachyarrhythmia among the leading causes of death in patients taking ICI. [43, 19, 36, 40]

The recommended treatment for ICI-induced myocarditis is high-dose corticosteroids such as methylprednisolone. Steroid use is based on its anti-inflammatory action and decreasing inflammation size. In addition to corticosteroid, immunosuppressive agents as infliximab [15, 17, 18, 31, 37, 38], intravenous immunoglobulin (IVG) [11, 13, 22, 24, 37, 38, 41], antithymocyte [35], and plasma exchange [19, 24, 33], and plasmapheresis [36, 38] are used as a secondary treatment after corticosteroid trial. [11] Also, myco-phenolate mofetil (MMF) [27, 28, 30] and tacrolimus [11, 20, 24, 41], tofacitinib [41], and abatacept [32] were used and they helped in the management and controlling the inflammation. The rationale for using infliximab and anti-TNF drug based on IC-induced myocarditis microscopic finding, these drugs decrease T-lymphocyte activity and have been considered for the treatment of myocarditis related to autoimmune or infectious disease, therefore, it is recommended to be used in IC-induced myocarditis.[4]

The current guideline for ICI-induced myocarditis treatment is based on expert consensus, not on clinical trial evidence-based, the recommendations state discontinuation of ICI, immunosuppressive with use of high dose steroid and heart failure treatment or arrhythmia treatment. [uptodate] We aim to assess the effectiveness of steroids in ICI-induced myocarditis and our results showed, with the use of steroids, the fatality in the 50 reviewed cases was 28%, and 64.15% of patients survived. In terms of steroid effectiveness 47.16% of patients improved while 52.83% did not improve completely and needed adding additional regimen, the commonly used added treatment infliximab, IgG, mycophenolate, Tacrolimus, plasma exchange, and plasmapheresis. Our reported fatality rate is about half what Matzen et al observed 48%. [10] The difference could be explained by the difference in inclusion and exclusion criteria between our reviews, we included only cases whose diagnoses were confirmed either by biopsy, CMRI or autopsy to minimize any confoundings. [Table 2] While Matzen et al did not specify that, therefore, it can’t be certain that all fatality cases were a result of ICI cardiac adverse events.

The causes of death in the 14th mortality were as following myocardial infarction (MI) [4, 18, 23, 33], multiple organ failure (MOF) [18, 36], complete heart block (CHB) [19, 36], ventricular tachycardia (VT) [18, 40], congestive heart failure (CHF) [19, 30], PE/bacteremia [31], hypotension/diaphragmatic paralysis [37], and progressive lung cancer. [11] The follow-up time ranged from 3 days to 26 months. 3 out of the 4 MI deaths occurred within 7 days of hospital admission, VT also within the first couple days of admission, lastly, the 2 deaths of CHB were on the third and 17th days. This should prompt the clinicians to be highly vigilant for acute deterioration of myocarditis cases and fatal complications that are associated with. One of the cases had myocarditis resolved and was discharged and died from MI in the 26 months from myocarditis diagnosis. One out of the 14 deceased did not receive steroids [40], 3 had infliximab [17, 18, 37] as an additional treatment, 2 IVG [11, 37], and 2 plasma exchange/plasmapheresis [19, 36]. Mehta et al concluded that myocarditis induced by ICI is a fatal complication of irAE steroid, alemtuzumab or abatacept are all possible treatments for IC-induced myocarditis, while infliximab is associated with increased risk of death from cardiovascular causes and should be avoided. [26] Dolladille et al argued against the current recommendation which states discontinuation of ICI therapy in life-threatening situations but Dolladille et al prefer management should be individualized or personalized cases by case based on the patient’s cancer status and toxicity progression and recommended both cardiological and oncological care approach. [44] Also, Huerta et recommended multidisciplinary teamwork including an oncologist, cardiologist, radiologist, and pathologist in the care of ICI patients to optimally manage and reduce mortality [4]

## Conclusion

Immune checkpoint inhibitors (ICI) are the therapy used against malignant neoplasms with or without combined drugs, so we are focusing on an adverse effect, myocarditis. This study shows that corticosteroids achieve a low mortality rate in patients with ICI-induced myocarditis, which, although it is a rare presentation, is serious. But the effectiveness of steroids was not high, the majority of patients needed another supplementary handling.

## Data Availability

All data produced in the present work are contained in the manuscript

## Acknowledgement

Mohammed Stohy, MBBCh and Madiha Zaidi, MBBS: participated in article screening and data extraction.

